# Patient Phenotyping for Atopic Dermatitis with Transformers and Machine Learning

**DOI:** 10.1101/2023.08.25.23294636

**Authors:** Andrew Wang, Rachel Fulton, Sy Hwang, David J. Margolis, Danielle L. Mowery

## Abstract

**Background:** Atopic dermatitis (AD) is a chronic skin condition that millions of people around the world live with each day. Performing research studies into identifying the causes and treatment for this disease has great potential to provide benefit for these individuals. However, AD clinical trial recruitment is a non-trivial task due to variance in diagnostic precision and phenotypic definitions leveraged by different clinicians as well as time spent finding, recruiting, and enrolling patients by clinicians to become study subjects. Thus, there is a need for automatic and effective patient phenotyping for cohort recruitment.

**Objective:** Our study aims to present an approach for identifying patients whose electronic health records suggest that they may have AD.

**Methods:** We created a vectorized representation of each patient and trained various supervised machine learning methods to classify when a patient has AD. Each patient is represented by a vector of either probabilities or binary values where each value indicates whether they meet a different criteria for AD diagnosis. Results: The most accurate AD classifier performed with a class-balanced accuracy of 0.8036, a precision of 0.8400, and a recall of 0.7500 when using XGBoost (Extreme Gradient Boosting).

**Conclusions:** Creating an automated approach for identifying patient cohorts has the potential to accelerate, standardize, and automate the process of patient recruitment for AD studies; therefore, reducing clinician burden and informing knowledge discovery of better treatment options for AD.

## Introduction

### Background

Atopic dermatitis (AD) is a common skin disease with a population prevalence of approximately 30% [1]. It is often diagnosed in early childhood, but onset can occur at any age [2–5]. Symptoms of AD include inflamed, red, irritated, and itchy skin and can cause significant physical and emotional distress. AD is often associated with other allergic illnesses including asthma, seasonal allergies, and food allergies [2,3,5–7].

AD is thought to be associated with skin barrier dysfunction and immune dysregulation [5]. AD has also been associated with genetic variation as well as environmental factors [5]. Classic treatment for AD has included the use of moisturizers, topical steroids, and other topical anti-inflammatory agents [8]. However, in the past few years, there have been significant treatment advances, which include systemic agents that alter immune function such as dupilumab. Therefore, due to the widespread nature of AD, the need for improved knowledge of the natural history of AD, the need to understand the efficacy of new treatments, and the need to develop new treatments, there is an urgent need to understand the clinical course of individuals with AD. However, identifying appropriate cohorts of patients for medical studies can be difficult and time consuming. Because AD is so common as well as diagnosed and managed by many different clinicians in varying healthcare settings, a potential source population would be patients from a health system’s electronic health records (EHRs) [9]. Investigators often ascertain a patient’s illness using International Classification of Disease (ICD) hospital billing codes as recorded during routine office visits. However, it has been previously demonstrated that reliance on ICD codes is not an accurate method for the ascertainment of AD study cohorts [9,10]. Furthermore, epidemiologic studies have used different methods and algorithms including the UK Working Party (UKWP) diagnostic criteria and the Hanifin and Rajka (HR) criteria [11,12]. Investigators attempting to conduct clinical trials and observational studies have also relied on manual, large-scale chart review, a process that is inefficient, slow, and tedious [9]. This motivates the need for a standard method to accurately, automatically, and efficiently identify potential patient cohorts from their text medical records via natural language processing (NLP) and machine learning (ML) techniques.

### Prior Work

Previously, researchers have aimed to phenotype patients with AD using EHR data. In particular, Gustafson et al. trained a logistic regression model with lasso regularization to identify cases of AD from the Northwestern Medical Enterprise Data Warehouse (NMEDW) which contained both structured data (ICD-9/ICD-10 codes, medication prescriptions, and lab results), as well as unstructured data (clinician notes from patient encounters) [10]. A gold standard diagnosis was assigned to each patient in their dataset by two rheumatologists following a chart review when using the UK Working Party (UKWP) criteria, and (alternatively) when using the Hanifin and Rajka (HR) criteria.

Although similar, our work differs in the following ways: 1) we survey a wide range of supervised machine learning algorithms as opposed to only using lasso regularized logistic regression, 2) we use transformer embeddings of sentences to represent information in each patient’s records and aggregate these embeddings with MLP networks to create a patient vector representation for patient phenotyping, and 3) we performed an ablation study of processing methods to compare the impact on performance in using a probability-based vs binary label of whether each patient meets various AD diagnostic criteria when creating a vector to represent each patient for input to our final AD patient phenotyping algorithms.

### Contributions

The primary contributions of our paper are as follows:

- We introduce and validate a rules-based approach for aggregating information from patient electronic health record (EHR) data to generate binary-valued patient vectors that are used with standard ML algorithms for patient phenotyping
- We introduce and validate a transformer-based approach for aggregating information and patient phenotyping by using BERT models (BERT Base Uncased and BioClinical BERT) to generate patient vectors of probabilities, which are used with standard ML algorithms for patient phenotyping.
- We compare the aforementioned approaches to 1) discern whether a transformer model pretrained on clinical text can provide performance benefits over a transformer model not pretrained on clinical text, and to 2) discern whether a transformer-based approach for aggregating information could outperform a rules-based approach for aggregating information.
- We demonstrate that multi-layer perceptron (MLP) networks can be used with BERT sentence embeddings to identify which sentences in patient records are relevant to the diagnosis of atopic dermatitis. These MLP networks can then be used during clinician chart review to highlight sentences that are relevant to diagnosis and therefore accelerate the process of chart review during clinical trial recruitment.

## Methods

### Overview

To predict whether a patient may qualify as a subject for an AD study based on their electronic health record, we first assigned patients in our dataset to either the training or testing sets. Then, for each patient, we aggregated the text from their EHR and constructed a vector representation of clinical features indicative of AD according to the UKWP criteria. Lastly, we leveraged our vectorized patient representations to train several machine learning classifiers to predict whether each patient has AD. In the following sections, we detail this process.

### Dataset Creation

We initially sampled 2,000 patients and their clinical records from Epic Clarity, Penn Medicine’s EHR database. We selected Penn Medicine patients who were diagnosed with a subset of AD-related ICD codes [9]. Of the 2,000 sampled patients, we identified 1,926 patients who had clinical notes for processing. We then de-identified these patient records according to the Safe Harbor method using PHIlter [13]. Each patient in the dataset was also manually reviewed and labeled according to the UK Working Party (UKWP) diagnostic criteria for AD. According to the UKWP criteria, in order to qualify as having AD, a patient must have an itchy skin condition along with 3 or more of the following: a history of flexural involvement, a history of asthma or hay fever, a history of dry skin, an onset of rash under the age of 2 years, or a visible flexural dermatitis. Our dataset was validated by two clinicians (a board-certified dermatologist (DJM) and a medical fellow (RF)), resulting in 137 patients with AD and 1,789 patients without AD.

**Figure 1.**
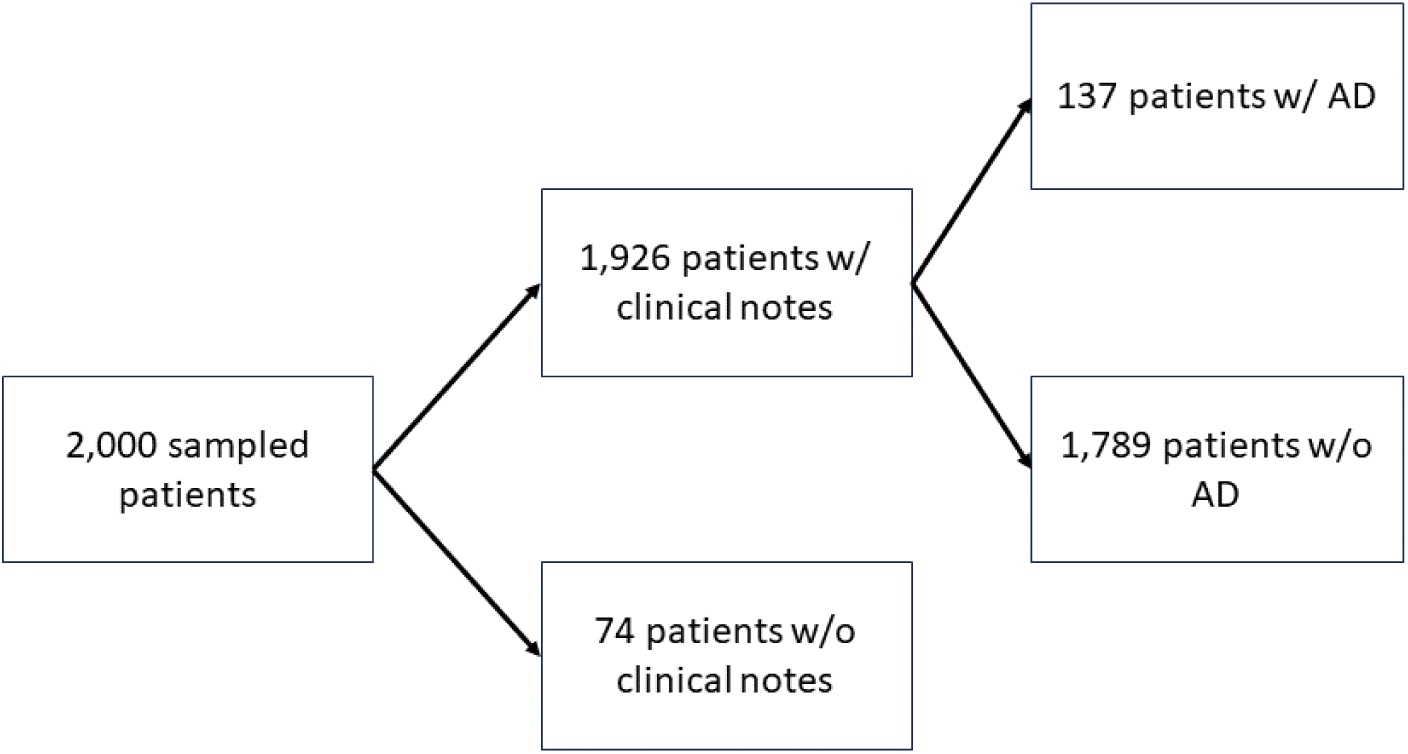
Waterfall diagram of cohort

### Training and Testing Split

We first created our training set. Due to the heavy class imbalance in our dataset, we decided to create a balanced training set to prevent biasing the model towards either AD or non-AD patients. In particular, we created the training set by assigning 80% of the 137 patients with AD to our training set, and under sampling from the non-AD patients to match the number of patients with AD. The remaining 20% of the 137 patients were assigned to both of our testing sets. This resulted in a training set that had 109 patients with AD, and 109 patients without AD.

Next, we created two testing sets. The first testing set was class-balanced and was intended to show how our patient classification model can generalize to unseen samples if the class distribution was kept the same. The second testing set was class-imbalanced (30% of patients with AD and 70% of patients without AD), and was intended to show how our patient classification model can perform when the class-distribution of the dataset matches the prevalence of AD in the United States.

We created the first (balanced) testing set by including the 20% (previously reserved for testing) of the 137 patients with AD, and combining them with an equal number of patients without AD who have not been used during training. This resulted in a (balanced) testing set that had 28 patients with AD and 28 patients without AD.

Furthermore, we created the second (unbalanced) testing set by including the same 20% of the 137 patients with AD, but instead combining them with a greater number of patients without AD to match the 30% prevalence rate of AD found in the US [1]. This resulted in a (unbalanced) testing set with 28 patients who have AD and 63 patients who don’t have AD.

We chose not to create a separate hyperparameter tuning set and instead applied cross validation for hyperparameter tuning on the training set due to the data scarce setting of our experiments.

### Vector Representation for AD Classification

Next, we created a vector representation for each patient. We performed 3 experiments to compare different methods of creating each patient’s vector representation (Figure 2).

**Figure 2.**
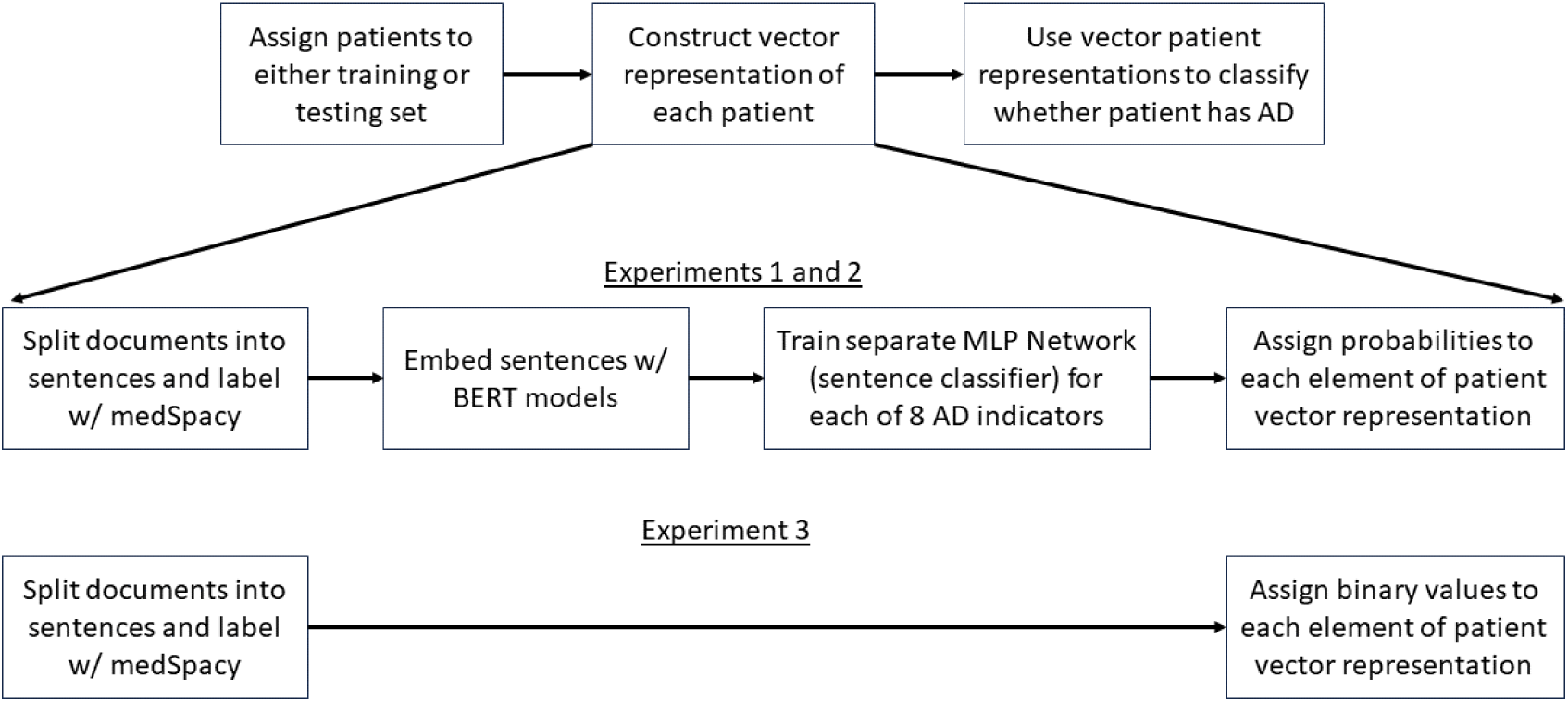
AD Phenotyping pipeline across all 3 experiments

#### Description of Patient Vector Representation

Each patient’s vector representation is 8 elements long, where each element of the vector is representative of whether the patient fulfills a different AD diagnosis criteria based on the UKWP criteria as well as clinician feedback (Table 1). Across all three experiments, each element in the patient vector corresponds to a distinct classification task; however in experiments 1 and 2 each element is a probability, and in experiment 3 each element is a binary value.

**Table 1.** Meaning of each patient vector element.

| Element | AD Indicator (Diagnostic Criteria) |
| --- | --- |
| 1 | EHR directly mentions patient has AD |
| 2 | Patient has hay fever allergies |
| 3 | Patient has atopic allergies |
| 4 | Patient has eczema or rashes |
| 5 | Patient has dry or itchy skin |
| 6 | Patient uses non-asthma medications related to treating AD |
| 7 | Patient has asthma |
| 8 | Patient uses asthma medications |

In experiments 1 and 2, elements 1 through 8 of each patient’s vector represent the highest probability that any sentence in the patient’s EHR mentions 1) AD or synonyms of AD, 2) keywords that suggest hay fever allergies, 3) keywords that suggest atopic allergies, 4) keywords that suggest eczema or rashes, 5) keywords that indicate dry or itchy skin, 6) keywords denoting non-asthma medications, 7) keywords suggesting the presence of asthma, and 8) keywords indicating the use of asthma medications.

In experiment 3, instead of each element representing a probability, each element represents a binary value of whether there was at least 1 sentence in the corresponding patient record suggesting the presence of the corresponding AD indicator.

In the first two experiments, each patient’s vector elements represent probabilities (ranging from 0 to 1). Each probability value is derived from a distinct MLP classifier. Experiments 1 and 2 were performed to compare the use of two BERT models (BERT Base Uncased [14,15] in experiment 1, and BioClinical BERT [16,17] in experiment 2) for creating sentence embeddings used to train multi-layer perceptron (MLP) networks (or alternatively, sentence classifiers). A separate MLP network is trained for each element of the patient vector. Each MLP network is trained to distinguish sentences in one of the 8 AD indicator categories from sentences in all other categories. Furthermore, medSpacy was used to split documents into sentences and label each sentence with different categories. After each sentence classifier is trained, embeddings of all sentences in each patient’s full EHR are passed through each sentence classifier and an aggregation function (max operator) is used to assign a value to each element of each patient’s vector. Our goal in experiments 1 and 2, was to test the hypothesis that a BERT model pretrained on clinical text (BioClinical BERT) could outperform a BERT model trained on non-clinical text (BERT Base Uncased).

In experiment 3, each patient’s vector elements are binary (either 0 or 1). Each element corresponds to a diagnostic criteria and represents whether medSpacy was able to identify at least 1 sentence in the patient’s record with a keyword and affirming context that suggests the patient meets the corresponding diagnostic criteria. Our goal was to conduct an ablation study to test the hypothesis that an AD patient classifier leveraging BERT embeddings to create the patient vector representation will better discern whether a patient has AD than an AD patient classifier without BERT embeddings.

#### Preprocessing for Experiments 1-3

Before each experiment, we applied the same preprocessing steps to assign one or more labels to each sentence in our corpus of documents in both our training and testing sets. Each sentence can be labeled as applying to one, multiple, or none of the 8 AD indicators previously defined.

For each of the 8 diagnostic criteria, we first created a list of keywords and phrases (for each vector element) that suggested the presence of the corresponding diagnostic criteria. Next, we used medSpacy with the ConText algorithm to split each document into sentences and categorize each sentence [18]. Using medSpacy allows us to obtain sentences that suggest the presence of each of the 8 diagnostic criteria due to medSpacy’s use of regex and rules-based keyword matching. Furthermore, medSpacy’s implementation of the ConText algorithm allows us to discern between sentences that affirm from negated assertions. We define negated sentences for each AD indicator as sentences where the indicator is ruled out, sentences where the indicator is experienced by someone other than the patient, and sentences where the existence of the indicator is hypothetical [19–22].

After assigning one or more categorical labels to each sentence with medSpacy, we then performed 3 different experiments to create a vectorized representation of each patient.

In tables 2 and 3 below, we include some statistics on the dataset obtained after preprocessing.

**Table 2.** Differences in number of documents, sentences, and tokens between AD and non-AD patients.

|  | AD Patient | AD Patient | Non-AD Patient | Non-AD Patient |
| --- | --- | --- | --- | --- |
|  | BERT Uncased | BioClinical BERT | BERT Uncased | BioClinical BERT |
| Avg # docs (per patient) | 23.44 | 23.44 | 7.99 | 7.99 |
| Avg # sentences (per patient) | 392.99 | 392.99 | 193.69 | 193.69 |
| Avg # tokens (per patient) | 16035.39 | 17054.11 | 7241.02 | 7674.35 |
| Avg # sentences (per doc) | 16.77 | 16.77 | 24.25 | 24.25 |
| Avg # tokens (per doc) | 684.16 | 727.63 | 906.45 | 960.69 |
| Avg # tokens (per sentence) | 40.80 | 43.40 | 37.38 | 39.62 |

**Table 3.**
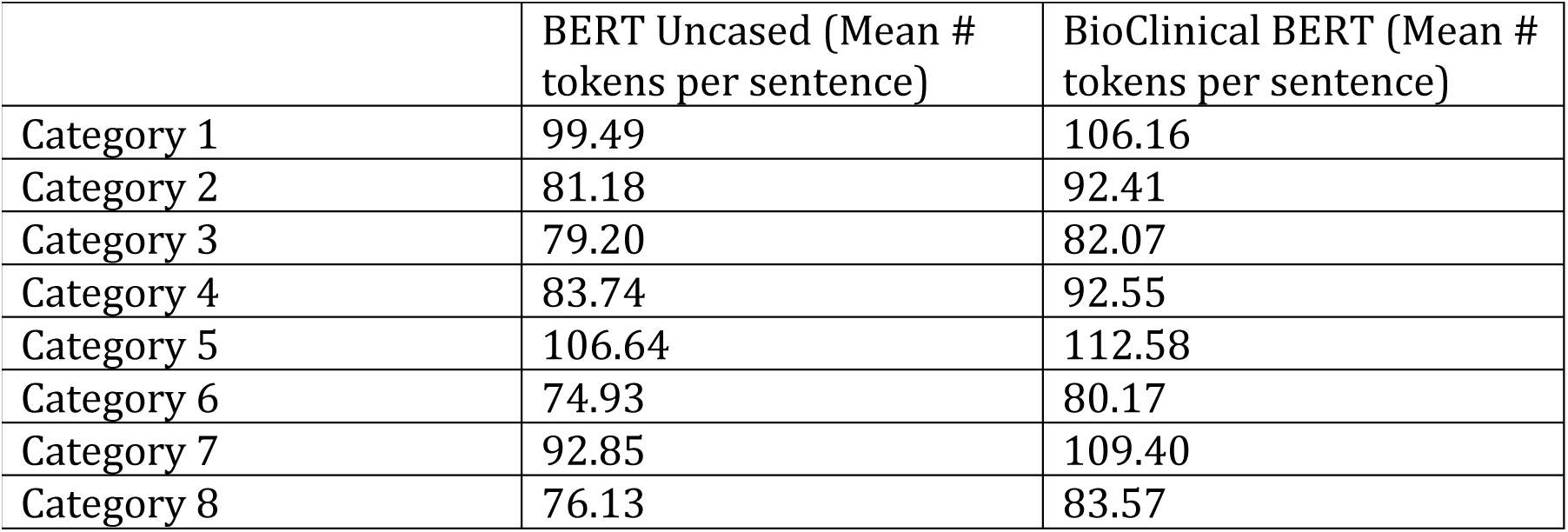
Mean number of tokens for sentences identified in each category.

As shown in table 2, AD patients have approximately twice as many sentences as non-AD patients. The average number of documents and sentences are the same (within patients who have AD, and similarly within non-AD patients) between BERT Base Uncased and BioClinical BERT experiments because these values are only dependent on medSpacy’s preprocessing of documents. Furthermore, using BioClinical BERT to tokenize sentences tends to yield more tokens (on average) per patient and per document. We hypothesize this is because the BioClinical BERT tokenizer is able to recognize more clinical terms and therefore yields more tokens for the same sentence than using the tokenizer from BERT Base Uncased.

As shown in table 3, sentences in category 5 (relating to dry or itchy skin) tend to have the most tokens, whereas sentences in category 6 (relating to the use of non-asthma medications related to treating AD) tend to have the least number of tokens. We hypothesize that this is because categories where the average number of tokens per sentence is greater tend to correspond to more general categories where many terms and sentences could apply, whereas categories where the average number of tokens per sentence is lower tend to correspond to more specific categories thus yielding a lower average number of tokens per sentence. Additionally, similarly to before, we can see that using BioClinical BERT tends to result in a greater number of tokens per sentence than using BERT Base Uncased for the same sentence.

#### Experiments 1 and 2 – Patient Vector Construction with BERT Embeddings

In experiments 1 and 2, we first used the sentences medSpacy identified in each category to create class-balanced training and testing sets for each MLP network classifier, as shown in table 4. The same training and testing set was used for both experiment 1 (BioClinical BERT) and experiment 2 (BERT Base Uncased).

**Table 4.** Training and testing dataset size for each classifier.

| Classifier | # of training samples | # of testing samples |
| --- | --- | --- |
| 1 | 2766 | 862 |
| 2 | 1302 | 392 |
| 3 | 532 | 168 |
| 4 | 9822 | 2454 |
| 5 | 1466 | 354 |
| 6 | 9114 | 2316 |
| 7 | 1596 | 520 |
| 8 | 4764 | 1070 |

Next, we used pretrained BERT models to generate embeddings of the sentences in each classifier’s training and testing set. We incorporated pretrained BERT models because these models have been trained on a much larger corpus than our existing dataset, and BERT provides a context sensitive embedding of text which other techniques such as bag of words don’t provide. Furthermore, we used BERT Base Uncased in experiment 1, and Alsentzer et. al’s BioClinical BERT in experiment 2 because we wanted to quantify how much of a difference in performance that using a model pretrained on clinical text can provide over a model that has not been pretrained on clinical text.

Using these embeddings, we trained a multi-layer perceptron (MLP) network to distinguish sentence embeddings in each category from sentence embeddings that aren’t in the corresponding category. Each of our MLP’s were trained with the following architecture: a fully connected input layer of shape 768 by 100, followed by a ReLU (Rectified Linear Unit) activation, further followed by a fully connected output layer of shape 100 by 2. We trained each of our MLP’s for 10 epochs with the cross-entropy loss function, the stochastic gradient descent (SGD) optimizer, a learning rate of 0.001, and a momentum value of 0.9. The final layer of each MLP can then be used to obtain the probability that any given sentence embedding comes from the category for which the MLP is being trained by passing the logits of the final layer to the softmax function.

We used the Rectified Linear Unit activation function as defined below, where x is the input to the ReLU function:

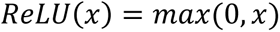

We also used the softmax function as defined below, where e is the standard exponential function, 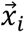 is element at index i of the K element long input vector 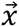.

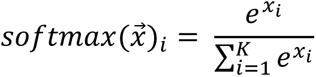

We chose to embed our sentences once with pretrained BERT models, and then feed these saved embeddings to our MLP networks as opposed to adding a classification head (a linear layer) to the end of our pretrained BERT models. Although doing so only allows us to fine tune the weights in our MLP network (as opposed to also fine tuning the weights BERT uses to embed the sentences), doing so allows us to iterate over different experiments more quickly and with less computational power. In particular, we are able to 1) avoid the large computational expense of gradient calculations during backpropagation for all 12 layers of transformers used by BERT when fine tuning the model, 2) avoid the computational expense of repeatedly generating the same embeddings from BERT multiple times (if we chose to freeze the weights of BERT and only fine tune an added classification head/linear layer), and 3) iterate more efficiently over different hyperparameter combinations across different experiments with our MLP networks.

**Figure 3.**
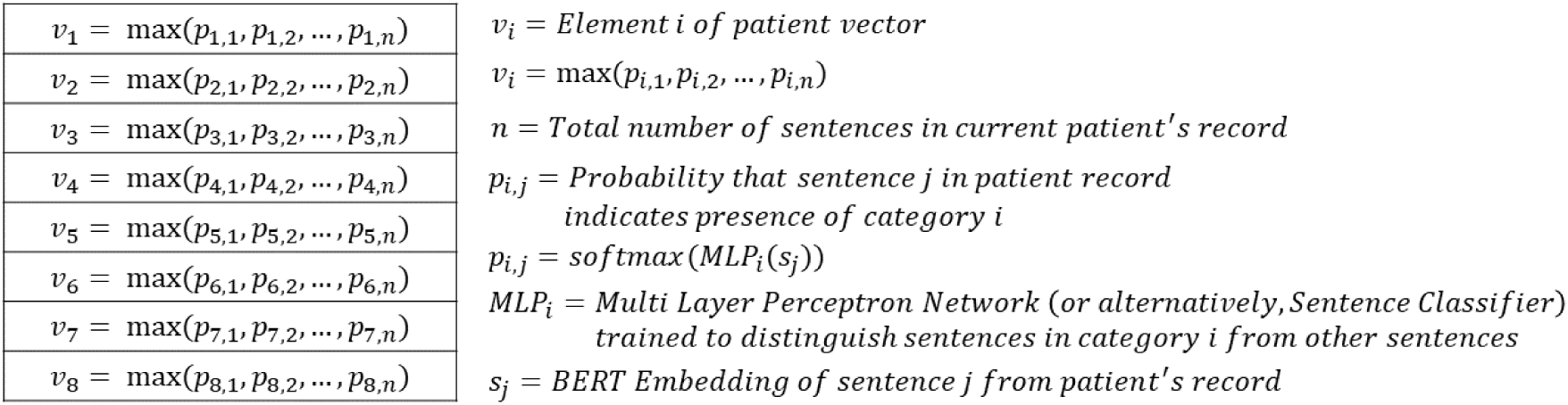
Patient vector representations of AD indicators in experiments 1 and 2

After training a separate MLP network for each of the 8 categories, we generated a vector representation for each patient where each of the 8 vector elements represents the highest probability that any given sentence in the patient record affirms the presence of the corresponding AD indicator. We accomplished this by iterating through all sentences in each patient’s full EHR and passing the sentence embedding through each of our 8 trained MLP networks to obtain 8 probabilities for each sentence corresponding to the probability that the sentence affirms each of the 8 AD indicators we previously selected. Then, for each patient and for each AD indicator, we kept the highest probability that any given sentence in the patient’s record affirms the presence of the AD indicator.

#### Experiment 3 – Patient Vector Construction without BERT Embeddings

In experiment 3, we generated each patient’s vector representation by assigning a 1 to each element of the patient vector if medSpacy with the ConText algorithm identified at least 1 sentence in the patient’s record that affirms or suggests the presence of the AD indicator for which the vector element corresponds to. Experiment 3 was conducted as an ablation study to quantify the performance benefit (if at all) of using contextual BERT text embeddings to generate probability scores that the patient meets various AD indicators.

**Figure 4.**
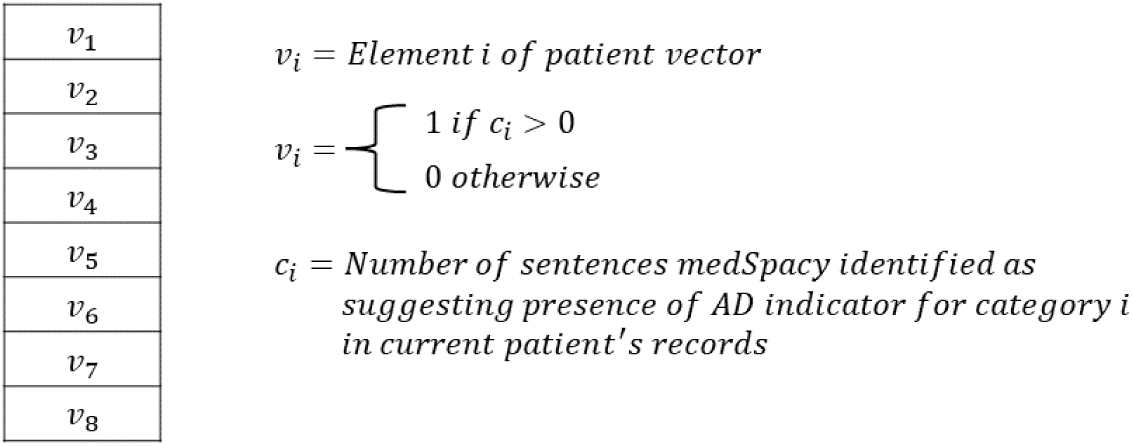
Patient vector representations of AD indicators in experiment 3

### AD Phenotyping with Vector Representations

In all three experiments, after generating a vector representation for each patient, we collated each patient vector representation with the corresponding label our clinicians assigned the patient when validating the dataset. Then, we fed the vector patient representation and corresponding patient label through a variety of classification algorithms. These include logistic regression, support vector machines (SVM), decision trees, random forests, K nearest neighbors (KNN), Extreme Gradient Boosting (XGBoost), and Adaptive Boosting (AdaBoost). During training for each of the previously mentioned classifiers, we used 5-fold cross validation to determine the best set of hyperparameters to use (as opposed to creating a separate validation set) due to the data scarce setting of our experiments. We then used the selected hyperparameters to train each algorithm on the entire training set and evaluated performance on the unbalanced and balanced testing sets. In addition to using the previously mentioned classifiers, we also used the stacking algorithm provided by scikit-learn to obtain an ensemble prediction from the different classifiers [23]. To quantify performance, we calculated the accuracy, precision, recall, F1-score, negative predictive value (NPV), and specificity of each algorithm on both testing sets.

We define accuracy, precision, and recall as follows, where TP is the number of true positives, TN is the number of true negatives, FP is the number of false positives, and FN is the number of false negatives:

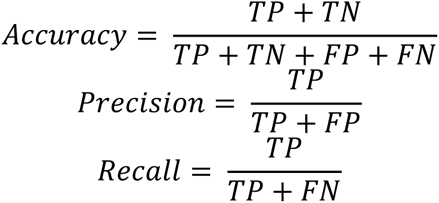

Additionally, we define the F1-score, NPV, and specificity as follows:

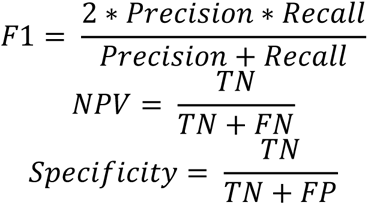

## Results

### Performance of MLP Networks

In this section, we compare the performance of several MLP classifiers in distinguishing sentences relevant to diagnosis of AD. This corresponds to the “Train separate MLP network (sentence classifier)” box in Figure 2.

As part of our AD Phenotyping pipeline, we trained various MLP networks to classify when a given sentence embedding indicates the presence of an AD indicator, and we compared performance of BioClinical BERT embeddings to BERT Base Uncased embeddings when training these MLP networks. In both cases, the classifier with the highest accuracy was the classifier for category 1 (sentences with direct mentions of AD). The classifiers with the two lowest accuracies were either the classifier for category 5 (sentences with mentions of dry or itchy skin) or the classifier for category 7 (sentences with mentions of asthma) for both the use of BioClinical BERT embeddings and the use of BERT Base Uncased embeddings.

However, the accuracy in classifier 7 was lower when using BERT Base Uncased embeddings than when using BioClinical BERT embeddings.

In experiment 1, the accuracies across AD indicator classifiers ranged from 0.7373 (classifier 5) to 0.9002 (classifier 1) as shown in table 5 below.

**Table 5.** Accuracy of different multi-layer perceptron networks in discerning sentences by AD indicator categories using BioClinical BERT sentence embeddings.

| Classifier | AD Indicator | Accuracy |
| --- | --- | --- |
| 1 | Direct mention of AD | 0.9002 |
| 2 | Mention of hay fever allergies | 0.8954 |
| 3 | Mention of atopic allergies | 0.8214 |
| 4 | Mention of eczema or rash | 0.8284 |
| 5 | Mention of dry or itchy skin | 0.7373 |
| 6 | Mention of non-asthma medications | 0.8204 |
| 7 | Mention of asthma | 0.7712 |
| 8 | Mention of asthma medications | 0.8299 |

In experiment 2, the accuracies across AD indicator classifiers ranged from 0.7269 (classifier 7) to 0.9153 (classifier 1) as shown in table 6 below.

**Table 6.** Accuracy of different multi-layer perceptron networks in discerning sentences by AD indicator categories using BERT Base Uncased sentence embeddings.

| Classifier | AD Indicator | Accuracy |
| --- | --- | --- |
| 1 | Direct mention of AD | 0.9153 |
| 2 | Mention of hay fever allergies | 0.7730 |
| 3 | Mention of atopic allergies | 0.7976 |
| 4 | Mention of eczema or rash | 0.8439 |
| 5 | Mention of dry or itchy skin | 0.7288 |
| 6 | Mention of non-asthma medications | 0.8096 |
| 7 | Mention of asthma | 0.7269 |
| 8 | Mention of asthma medications | 0.8738 |

### AD Phenotyping with Patient Vector Representations

In this section, we compare performance in patient classification when using different methods for creating patient vector representations. This encompasses all three experiments and corresponds to the “Use vector patient representations to classify whether patient has AD” box in Figure 2.

In experiment 1, we leveraged BioClinical BERT sentence embeddings to train various MLP networks to discern sentence embeddings in different AD indicator categories. Then, we applied these trained MLP networks (sentence classifiers) along with an aggregation function (max operator) to assign values to each element of each patient’s vector representation. Lastly, we used each patient’s vector representation with their validated label to train various ML algorithms. We evaluated on both a balanced and unbalanced testing set.

As shown in table 7, the accuracy on the balanced testing set ranges from 0.5893 (Decision Tree) to 0.7321 (Logistic Regression and SVM).

**Table 7:** AD Phenotyping Performance on Balanced Testing Set in Experiment 1 (BioClinical BERT)

| Model | Accuracy | Precision | Recall | F1 | NPV | Specificity |
| --- | --- | --- | --- | --- | --- | --- |
| Logistic Regression | 0.7321 | 0.7241 | 0.7500 | 0.7368 | 0.7407 | 0.7500 |
| SVM | 0.7321 | 0.7826 | 0.6429 | 0.7059 | 0.6970 | 0.7857 |
| Decision Tree | 0.5893 | 0.6316 | 0.4286 | 0.5106 | 0.5676 | 0.7500 |
| Random Forest | 0.6964 | 0.7037 | 0.6786 | 0.6909 | 0.6897 | 0.8214 |
| KNN | 0.6786 | 0.7273 | 0.5714 | 0.6400 | 0.6471 | 0.7857 |
| XGBoost | 0.6071 | 0.6154 | 0.5714 | 0.5926 | 0.6000 | 0.8571 |
| AdaBoost | 0.6429 | 0.6538 | 0.6071 | 0.6296 | 0.6333 | 0.7857 |
| Stacking Classifier | 0.6964 | 0.7391 | 0.6071 | 0.6667 | 0.6667 | 0.7500 |

As shown in table 8, the range of accuracies on the unbalanced testing set is slightly lower, ranging from 0.5824 (Decision Tree) to 0.7253 (Stacking Classifier).

**Table 8:** AD Phenotyping Performance on Unbalanced Testing Set in Experiment 1 (BioClinical BERT)

| Model | Accuracy | Precision | Recall | F1 | NPV | Specificity |
| --- | --- | --- | --- | --- | --- | --- |
| Logistic Regression | 0.6813 | 0.4884 | 0.7500 | 0.5915 | 0.8542 | 0.6984 |
| SVM | 0.6923 | 0.5000 | 0.6429 | 0.5625 | 0.8181 | 0.7302 |
| Decision Tree | 0.5824 | 0.3438 | 0.3929 | 0.3667 | 0.7119 | 0.7143 |
| Random Forest | 0.7143 | 0.5313 | 0.6071 | 0.5667 | 0.6845 | 0.7619 |
| KNN | 0.6593 | 0.4571 | 0.5714 | 0.5079 | 0.7857 | 0.7937 |
| XGBoost | 0.6264 | 0.4211 | 0.5714 | 0.4848 | 0.7736 | 0.7619 |
| AdaBoost | 0.6044 | 0.4048 | 0.6071 | 0.4857 | 0.7755 | 0.7302 |
| Stacking Classifier | 0.7253 | 0.5429 | 0.6786 | 0.6032 | 0.8393 | 0.6984 |

In experiment 2, we followed the same process as in experiment 1; however, we used BERT Base Uncased instead of BioClinical BERT. As shown in table 9, the accuracy of our AD patient classifiers on the balanced testing set ranges from 0.5179 (AdaBoost) to 0.6250 (Random Forest).

**Table 9:**
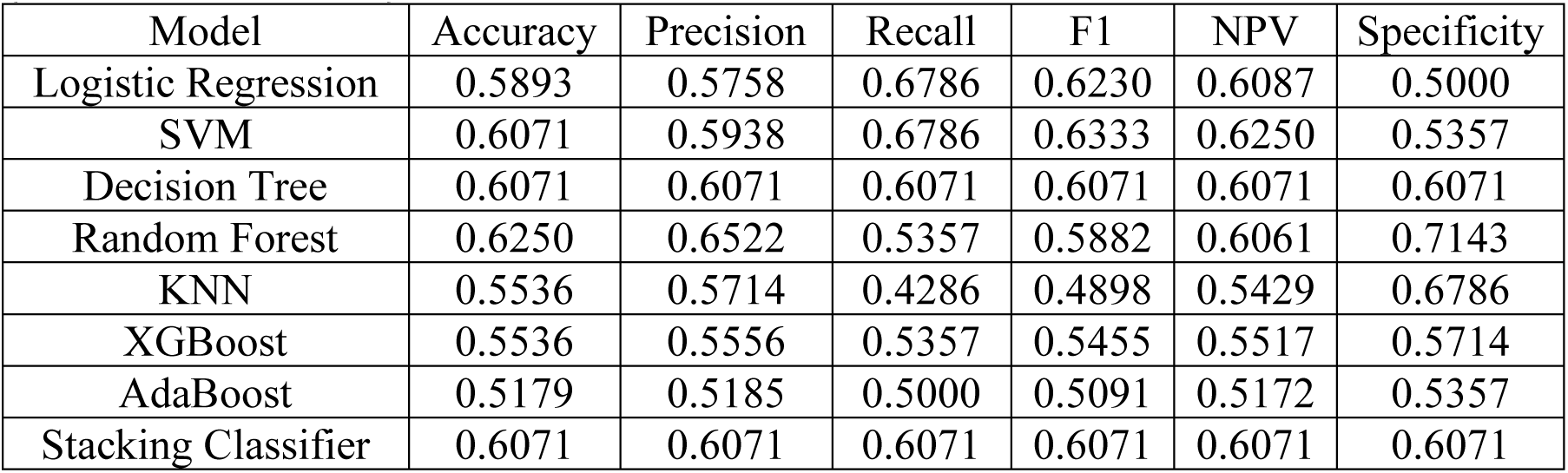
AD Phenotyping Performance on Balanced Testing Set in Experiment 2 (BERT Base Uncased)

As shown in table 10, the range of accuracies of our AD patient classifiers on the unbalanced testing set is slightly higher, ranging from 0.5714 (Logistic Regression and SVM) to 0.6703 (Random Forest).

**Table 10:** AD Phenotyping Performance on Unbalanced Testing Set in Experiment 2 (BERT Base Uncased)

| Model | Accuracy | Precision | Recall | F1 | NPV | Specificity |
| --- | --- | --- | --- | --- | --- | --- |
| Logistic Regression | 0.5714 | 0.3878 | 0.6786 | 0.4935 | 0.7857 | 0.5238 |
| SVM | 0.5714 | 0.3878 | 0.6786 | 0.4935 | 0.7857 | 0.5238 |
| Decision Tree | 0.6484 | 0.4474 | 0.6071 | 0.5152 | 0.7925 | 0.6667 |
| Random Forest | 0.6703 | 0.4737 | 0.6429 | 0.5455 | 0.8113 | 0.6825 |
| KNN | 0.6264 | 0.4000 | 0.4286 | 0.4138 | 0.7377 | 0.7143 |
| XGBoost | 0.6374 | 0.4286 | 0.5357 | 0.4762 | 0.7679 | 0.6825 |
| AdaBoost | 0.5934 | 0.3784 | 0.5000 | 0.4308 | 0.7407 | 0.6349 |
| Stacking Classifier | 0.6484 | 0.4474 | 0.6071 | 0.5152 | 0.7925 | 0.6667 |

In experiment 3, we performed an ablation study and assigned binary labels to the elements of each patient’s vector based on whether medSpacy was able to identify at least one sentence in each of the AD indicator categories that each vector element corresponds to. As shown in table 11, the accuracy across our AD patient classifiers on the balanced testing set ranges from 0.6964 (KNN) to 0.8036 (XGBoost).

**Table 11:** AD Phenotyping Performance on Balanced Testing Set in Experiment 3 (Binary Vector Encoding)

| Model | Accuracy | Precision | Recall | F1 | NPV | Specificity |
| --- | --- | --- | --- | --- | --- | --- |
| Logistic Regression | 0.7679 | 0.7586 | 0.7857 | 0.7719 | 0.7778 | 0.7500 |
| SVM | 0.7857 | 0.7857 | 0.7857 | 0.7857 | 0.7857 | 0.7857 |
| Decision Tree | 0.7857 | 0.7667 | 0.8214 | 0.7931 | 0.8077 | 0.7500 |
| Random Forest | 0.7857 | 0.8077 | 0.7500 | 0.7778 | 0.7667 | 0.8214 |
| KNN | 0.6964 | 0.7391 | 0.6071 | 0.6667 | 0.6667 | 0.7857 |
| XGBoost | 0.8036 | 0.8400 | 0.7500 | 0.7925 | 0.7742 | 0.8571 |
| AdaBoost | 0.7857 | 0.7857 | 0.7857 | 0.7857 | 0.7857 | 0.7857 |
| Stacking Classifier | 0.7500 | 0.7500 | 0.7500 | 0.7500 | 0.7500 | 0.7500 |

As shown in table 12, the lower bound of the range of accuracies across our AD patient classifiers on the unbalanced testing set is higher and the upper bound of the accuracies is lower. The accuracies on the unbalanced testing set ranges from 0.7143 (Stacking Classifier) to 0.7582 (Random Forest and Stacking Classifier).

**Table 12:**
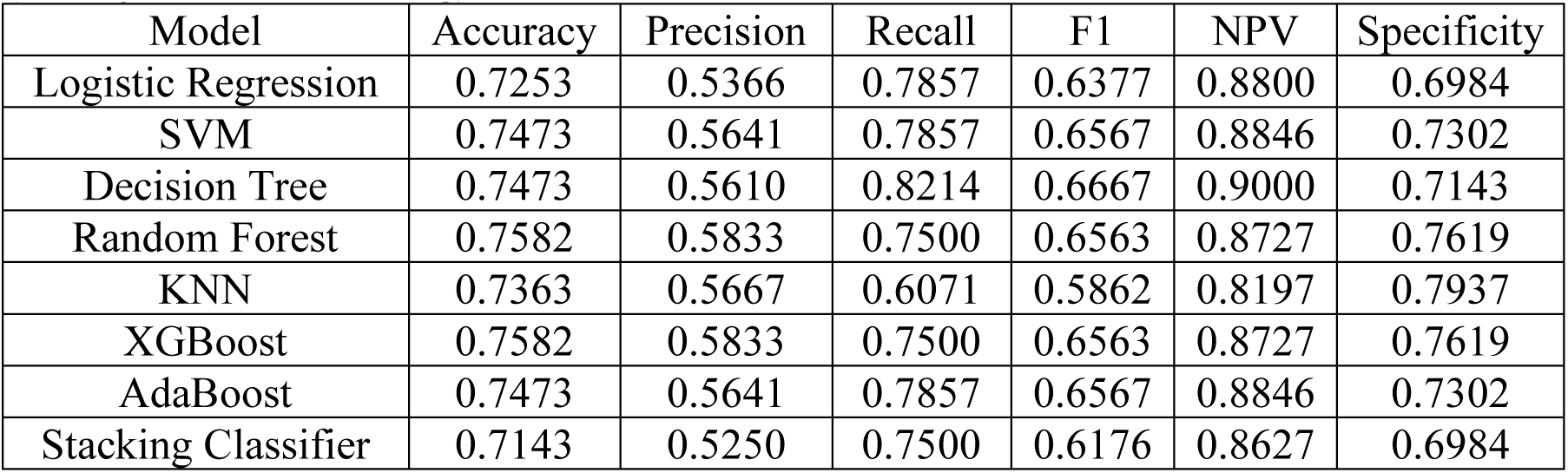
AD Phenotyping Performance on Unbalanced Testing Set in Experiment 3 (Binary Vector Encoding)

| Model | Accuracy | Precision | Recall | F1 | NPV | Specificity |
| --- | --- | --- | --- | --- | --- | --- |
| Logistic Regression | 0.7253 | 0.5366 | 0.7857 | 0.6377 | 0.8800 | 0.6984 |
| SVM | 0.7473 | 0.5641 | 0.7857 | 0.6567 | 0.8846 | 0.7302 |
| Decision Tree | 0.7473 | 0.5610 | 0.8214 | 0.6667 | 0.9000 | 0.7143 |
| Random Forest | 0.7582 | 0.5833 | 0.7500 | 0.6563 | 0.8727 | 0.7619 |
| KNN | 0.7363 | 0.5667 | 0.6071 | 0.5862 | 0.8197 | 0.7937 |
| XGBoost | 0.7582 | 0.5833 | 0.7500 | 0.6563 | 0.8727 | 0.7619 |
| AdaBoost | 0.7473 | 0.5641 | 0.7857 | 0.6567 | 0.8846 | 0.7302 |
| Stacking Classifier | 0.7143 | 0.5250 | 0.7500 | 0.6176 | 0.8627 | 0.6984 |

## Discussion

### Sentence Classification Results

We hypothesized that using BioClinical BERT sentence embeddings to train sentence classifiers would provide better performance than using BERT Base Uncased sentence embeddings due to the clinical setting of our data. Given the results in tables 5 and 6, we observed that this was most often true in the context of sentence classification because we were able to achieve better performance in the majority (5 out of 8) of the sentence classification tasks when using BioClinical BERT embeddings as opposed to BERT Base Uncased embeddings.

Using BioClinical BERT sentence embeddings yielded stronger performance when distinguishing sentences in 5 of the 8 sentence categories – category 2 (mentions of hay fever allergies), category 3 (mentions of atopic allergies), category 5 (mentions of dry or itchy skin), category 6 (mentions of non-asthma medications), and category 7 (mentions of asthma). More specifically, we observed higher accuracies when using BioClinical BERT sentence embeddings for classifiers 2 (0.8954), 3 (0.8214), 5 (0.7373), 6 (0.8204), and 7 (0.7712) than their corresponding counterparts when using BERT Base Uncased embeddings for classifiers 2 (0.7730), 3 (0.7976), 5 (0.7288), 6 (0.8096), and 7 (0.7269). We observed that the differences in performance between using BioClinical BERT embeddings and BERT Base Uncased embeddings are most pronounced for classifiers 2 and 7 which correspond to mentions of hay fever allergies and asthma mentions. We hypothesize this is because hay fever allergies and asthma (and their synonyms) may be very common terms in clinical notes; therefore, models trained on clinical data (BioClinical BERT) may be able to provide stronger performance than models trained on non-clinical text (BERT Base Uncased) which may not have as many mentions of hay fever allergies or asthma.

Conversely, using BERT Base Uncased embeddings yielded stronger performance when distinguishing sentences in the other 3 of 8 sentence categories – category 1 (direct mentions of AD), 4 (mentions of eczema or rashes), and 8 (mentions of asthma medications). More specifically, we observed higher accuracies when using BERT Base Uncased sentence embeddings for classifiers 1 (0.9153), 4 (0.8439), and 8 (0.8738) than their corresponding counterparts when using BioClinical BERT embeddings for classifiers 1 (0.9002), 4 (0.8284), and 8 (0.8299). We observed differences in performance between using BERT Base Uncased embeddings and BioClinical BERT embeddings are most evident for classifier 8 which corresponds to mentions of asthma medications. Although this is counterintuitive at first (we would expect that a classifier using embeddings generated from BioClinical BERT to be able to better recognize allergy medicines), we believe that the performance benefit from using BERT Base Uncased can be attributed to the list of terms we give to medSpacy when asking it to identify sentences in category 8. Many of the asthma medications in category 8 sentences are either monoclonal antibody medications ending in -mab (Ex: benralizumab, mepolizumab, omalizumab, etc) or hydrofluoroalkanes (Ex: atrovent hfa, flovent hfa, xopenex hfa, etc). Because monoclonal antibodies are very specialized types of medication, they may not occur as frequently as other terms in the corpus used to train BioClinical BERT so a more general model such as BERT Base Uncased may provide more robust performance. Additionally, because the hydrofluoroalkane allergy medications in category 8 sentences are often abbreviated with “hfa” which can have alternate medical meanings such as high-functioning autism or health facility administrator, the BioClinical BERT embeddings might not be representative of the presence of allergy medications in the sentence, so a more general model such as BERT Base Uncased may be able to provide better performance.

More broadly looking at the results in table 5 and 6, we can see that the least accurate classifier has an accuracy of 0.7288, while the most accurate classifier is able to achieve an accuracy of 0.9153. Furthermore, when aggregating the most accurate classifiers from both tables we can see that we are able to achieve accuracies of 0.9153 (classifier 1) for identifying sentences that directly suggest the patient has AD, 0.8954 (classifier 2) for identifying sentences that mention hay fever allergies, 0.8214 (classifier 3) for identifying sentences that mention atopic allergies, 0.8439 (classifier 4) for identifying sentences that mention eczema or skin rashes, 0.7373 (classifier 5) for identifying sentences that mention dry or itchy skin, 0.8204 (classifier 6) for identifying sentences that mention non-asthma medications related to diagnosis of AD, 0.7712 (classifier 7) for identifying sentences that mention asthma, and 0.8738 (classifier 8) for identifying sentences that mention asthma medications. Because our training and testing sets were both class-balanced and the majority (6 of the 8) of the most accurate classifiers previously mentioned achieve accuracies between 0.8204 and 0.9153, we believe these results are promising and indicate that our sentence classifiers could potentially be used to save time in a clinical setting during chart review by identifying (and highlighting for review) sentences relevant to diagnosis of AD when recruiting for clinical trials.

### AD Phenotyping Results

Per tables 7 through 10, our earlier hypothesis holds - using clinical embeddings (BioClinical BERT) to generate the patient vector representation does provide better performance in patient phenotyping than using non-clinical embeddings (BERT Base Uncased). Comparing evaluation on the balanced testing set in tables 7 and 9, we observe that using BioClinical BERT embeddings provides higher accuracy in almost all models, with the exception of Decision Trees where BERT Base Uncased provides better performance (accuracy of 0.6071) as compared to BioClinical BERT (accuracy of 0.5893). Comparing evaluation on the unbalanced testing set in tables 8 and 10, we observed that the same trend follows – using BioClinical BERT embeddings provides higher accuracy in almost all models, with the exception of Decision Trees and XGBoost where using BERT Base Uncased embeddings provides better performance (accuracy of 0.6484 for Decision Trees and 0.6374 for XGBoost) as compared to their counterparts with BioClinical BERT embeddings (accuracy of 0.5824 for Decision Trees and 0.6264 for XGBoost).

As part of our experimental design, we included an ablation study in experiment 3 so we could compare the difference in performance during patient phenotyping when removing the use of BERT models to create each patient’s vector representations. On the class-balanced testing set, we observed that accuracies range from 0.6071 to 0.7321 when using BioClinical BERT embeddings in table 7, accuracies range from 0.5179 to 0.6250 when using BERT Base Uncased embeddings in table 9, and accuracies range from 0.6964 to 0.8036 when removing the use of BERT models in table 11 (experiment 3). On the unbalanced testing set, we observed that accuracies range from 0.5824 to 0.7253 when using BioClinical BERT embeddings in table 8, accuracies range from 0.5714 to 0.6703 when using BERT Base Uncased embeddings in table 10, and accuracies range from 0.7143 to 0.7582 when removing the use of BERT models in table 12 (experiment 3).

In both cases (evaluation on the balanced testing set, and evaluation on the unbalanced testing set), we found that models in experiment 3 (our ablation study) generally outperform (or are as good as) their corresponding counterparts in experiments 1 and 2 (our BERT experiments) across all metrics (accuracy, precision, recall, F1, NPV, and specificity), with the exception that the stacking classifier in experiment 1 (BioClinical BERT) has marginally stronger accuracy and precision than the stacking classifier in experiment 3. This shows that traditional rules-based approaches (experiment 3) can outperform BERT-based approaches for generating a patient vector representation for downstream patient phenotyping.

We hypothesize that models in experiments 1 and 2 showed lower performance because errors from our sentence classifiers in earlier stages of the pipeline could have propagated to later stages of the pipeline during patient phenotyping. Because we leveraged the max operator to aggregate probabilities that any given sentence in the patient record applies to each category, more sentences in each patient record would lead to a greater chance that an erroneous prediction with a high probability would lead to a false positive error in the creation of each patient’s vector representation in experiments 1 and 2.

Although there is a wide range in performance for our AD patient phenotyping algorithms, we believe that we have reached our goal of developing a system capable of AD patient phenotyping for clinical trial recruitment because tables 11 and 12 show promising results. Furthermore, our system can be used as a first step during AD clinical trial recruitment to filter out most patients who may not qualify for AD trials and therefore save valuable clinician time. We believe our pipeline is important and valuable because unlike other diseases such as influenza, COVID-19, and cancer, there is no gold-standard test result that can be used to determine when a patient has atopic dermatitis. Instead, clinicians must spend large amounts of time undergoing chart review to individually determine whether each patient has atopic dermatitis.

### Limitations

One limitation of our study was the small size of our dataset. Although we had a total of 1,926 patients in our dataset, only 137 of them were validated as having AD. During training, we leveraged 109 of the 137 AD patients, and sampled another 109 non-AD patients to create a class balanced training set. The small size of the training set could lead to overfitting and therefore result in reduced performance on the testing set. Future work could involve obtaining more data from patients with AD, as well as exploring the use of an imbalanced dataset but using a class-weighted loss function to counteract the class-imbalance.

A second limitation of our study was the input limit size of the large language models that were used. Both BERT Base Uncased and BioClinical BERT had an input limit of 512 tokens. This meant that any input text that was longer than 512 tokens would be ignored when training BERT. Consequently, we couldn’t simply directly concatenate all documents from each patient’s EHR and feed the tokenized documents of each patient into BERT with an added classification head for training as well as direct prediction of whether the patient has AD. Instead, we designed a pipeline around distilling information from all documents in each patient’s EHR into a patient vector representation and then using this patient vector representation to train various classical ML algorithms for phenotyping the patient. Future work could involve exploring the use of other LLM’s that are suited for long inputs such as Longformer or Doc2Vec for predicting when a patient should be labeled as having AD.

A third limitation of our study was the list of AD indicators we selected. We didn’t consider additional AD indicators and we also did not consider the use of different combinations (or subsets) of the AD indicators selected. This is particularly relevant in considering that 1) our pipeline is intended to be used for identifying patients with AD, and 2) that one of our AD indicators (category 1) directly targets whether there is any given sentence in the patient’s record that mentions AD which could be in the context of a family history of AD, a potential (but not confirmed) diagnosis of AD, as well as a confirmed diagnosis of AD, among other possibilities. If this AD indicator is removed, then one interesting research question could be whether our pipeline is still able to maintain performance similarly to what it is currently able to achieve. Future work could involve assessing the performance impact from removing (or adding) the use of various AD indicators. We could then determine if our pipeline is relying too much on or overfitting to one or more indicators. Furthermore, we could also re-design our patient vector and separate the feature for category 1 (any sentence that mentions AD) into 3 separate indicators, whether there is 1) a family history of AD, 2) an affirmed diagnosis that the patient has AD, and 3) uncertainty of whether the patient has AD. Doing so could potentially improve precision.

### Potential Applications

Given the aforementioned results, we believe our AD classifier could be operationalized to facilitate reliable and efficient EHR chart review. For example, sentence classifiers could visually indicate AD indicators inline text, therefore reducing information foraging efforts by clinicians. Additionally, AD phenotyping classifiers could indicate the strength of a patient match to UKWP criteria, exact or partial, based on AD indicator sentence classifications. Furthermore, ranking patient cases by match strength could reduce the number of cases reviewed to generate both case and matched controls.

### Conclusions

In conclusion, we present and validate a promising pipeline for phenotyping patients with AD during clinical trial recruitment. To do so, we compare a rules-based and transformer-based approach for creating a vector representation of each patient, and compare downstream performance in patient phenotyping with various standard ML algorithms. We find that a traditional rules-based approach outperforms using a transformer-based approach (Experiment 3). We hope that our pipeline can be deployed in hospital settings during clinical trial recruitment as an initial first step to automatically filter candidates before manual review. Additionally, we show that multi-layer perceptron networks can identify whether sentences are relevant to AD diagnosis. These multi-layer perceptron networks can later be deployed in clinical settings to highlight which sentences relevant for physicians during manual chart review, therefore reducing physician burden. Future work can involve extending our patient phenotyping pipeline to other datasets and other diseases.

## Data Availability

To protect patient privacy, the clinical data is not available.

## Acknowledgements

AW designed the experiments, wrote the code, performed the experiments, wrote the first draft of the manuscript, and revised the manuscript.

DJM conceptualized and implemented the chart abstraction study, annotated the dataset, interpreted results, and revised the manuscript.

RF annotated the dataset and revised the manuscript.

SH queried and de-identified the dataset as well as revised the manuscript.

DLM conceptualized the study and experiment design, interpreted results, wrote and revised the manuscript, provided secure storage and compute resources.

This study was partially funded by the National Institutes of Health (NIH) National Institute of Arthritis and Musculoskeletal and Skin Diseases (NIAMS) P30-AR069589 as part of the Penn Skin Biology and Diseases Resource-Based Center (Core: DJM, DLM).

## Ethics Statement

This research protocol was reviewed and approved by the University of Pennsylvania Institute Review Board and determined as exempt (IRB#843922).

## Abbreviations

AD: atopic dermatitis
BERT: Bidirectional Encoder Representations from Transformers
EHR: Electronic Health Records
ICD: International Classification of Disease
UKWP: United Kingdom Working Party
HR: Hanifin and Rajka
AI: Artificial Intelligence
NLP: Natural Language Processing
ML: Machine Learning
MLP: Multi-layer Perceptron
ReLU: Rectified Linear Unit
SGD: Stochastic Gradient Descent
KNN: K-Nearest Neighbors
XGBoost: Extreme Gradient Boosting
AdaBoost: Adaptive Boosting
SVM: Support Vector Machines
TP: True Positive
TN: True Negative
FP: False Positive
FN: False Negative
NPV: Negative Predictive Value

